# Enhancing Recovery and Reducing Inflammation: The Impact of Enhanced Recovery After Surgery Recommendations on Inflammatory Markers in Laparoscopic Surgery – a Scoping review

**DOI:** 10.1101/2024.11.17.24317456

**Authors:** Carlos Darcy Alves Bersot, Lucas Ferreira Gomes Pereira, Victor Gabriel Vieira Goncho, José Eduardo Guimaraes Pereira, Luiz Fernando dos Reis Falcão

**Affiliations:** Department of Anesthesia, BP Hospital – A Beneficência Portuguesa de São Paulo (Anextesia), São Paulo, Brazil; Postgraduate in Translational Medicine of the Paulista School of Medicine, EPM-UNIFESP, São Paulo, Brazil; Discipline of Anesthesiology, Hospital das Clínicas da Faculdade de Medicina da Universidade de São Paulo, São Paulo, Brazil; Department of Anesthesiology, Hospital Unimed Volta Redonda, Rio de Janeiro, Brazil; Department of Anesthesiology, Pain and Critical Care Medicine, Federal University of São Paulo (EPM-UNIFESP), São Paulo, Brazil

**Keywords:** Enhanced Recovery After Surgery, Inflammation, Laparoscopy

## Abstract

**Introduction:** The relationship between the Enhanced Recovery After Surgery (ERAS) guidelines and inflammatory markers in laparoscopic surgery has garnered increasing attention. These recommendations are designed to minimize surgical stress and potentially improve recovery outcomes by modifying perioperative care.

**Objective:** This scoping review aims to evaluate the impact of ERAS recommendations on inflammatory markers in patients undergoing laparoscopic surgeries, identifying current research gaps and consolidating findings from existing studies.

**Methods:** Guided by the Cochrane Handbook for Systematic Reviews and adhering to the PRISMA-ScR guidelines, this review analyzed studies from databases like PubMed, Scopus, and Cochrane Library. We included both randomized controlled trials and observational studies that assessed inflammatory markers such as C-reactive protein (CRP), white blood cells (WBC), and Interleukin-6 (IL-6) in laparoscopic surgery patients managed with ERAS recommendations.

**Results:** Out of 64 initial studies, 7 met the inclusion criteria, involving a total of 2,047 patients. Most of the studies focused on laparoscopic colorectal surgeries. Commonly assessed markers were CRP and WBC. The findings consistently showed that ERAS guideline could mitigate the inflammatory response, evidenced by reduced levels of CRP and IL-6, which correlated with fewer postoperative complications and expedited recovery.

**Conclusion:** ERAS recommendations appear to beneficially modulate inflammatory responses in laparoscopic surgery, which suggests a potential for enhanced recovery outcomes. However, the evidence is currently limited by the small number of studies and inherent methodological biases. Further robust RCTs are required to strengthen the evidence base and refine these protocols for broader clinical application.

## INTRODUCTION

The association between inflammation and cancer has been extensively discussed since 1863, when Virchow observed that tumors frequently develop at sites of chronic inflammation.^(1)^ Research has consistently demonstrated that inflammation contributes to tumor growth and aggressiveness; both preoperative and early postoperative inflammatory responses can foster a micrometastatic environment and adversely affect cancer prognosis. ^(1-4)^

The initiative to reduce recovery times after surgery was pioneered in the USA under the concept of “fast-track” surgery, particularly aimed at expediting recovery following cardiac procedures.^(5)^ Kehlet et al. further advanced this concept by developing a multimodal rehabilitation program focused on colorectal surgeries, which was successful in reducing hospital stay durations.^(6)^ The Enhanced Recovery After Surgery (ERAS) programs have refined these preliminary concepts into a standardized, evidence-based approach that enhances surgical outcomes across various disciplines. Originating in Europe, ERAS Society unites diverse surgical teams dedicated to fostering comprehensive, multi-professional patient care.^(7)^

The cellular response to surgical tissue damage triggers the activation of macrophages and neutrophils within the innate immune system via the production of inflammatory cytokines, such as tumor necrosis factor (TNF) alpha, interleukin (IL)-1, and IL-6. These pro-inflammatory cytokines modify the levels of circulating acute-phase proteins, including C-reactive protein (CRP), albumin, ferritin, transferrin, and fibrinogen. ^(8)^ However, the pathophysiology of post-surgical recovery is not solely a consequence of tissue injury but is inherently multifactorial, encompassing elements such as anxiety, pain, coagulation disorders, hemodynamic changes, and hypoxia. Given the multitude of factors that promote inflammation during the surgical stress response, interventions proposed by the ERAS guidelines address these various components comprehensively.^(9)^

This scoping review aims to provide a descriptive summary of the studies included and to identify potential gaps in the literature regarding the impact of the ERAS guidelines on the inflammatory response following laparoscopic surgery. The guiding question for this review is: “What research has been conducted on the impact of the ERAS recommendations on inflammatory markers in laparoscopic surgery, and what evidence is available regarding its effects on the immune system?”

## METHODS

### Study Design and Protocol Registration

This scoping review was guided by the Cochrane Handbook for Intervention Reviews ^(10)^ and conducted in accordance with the Preferred Reporting Items for Systematic Reviews and Meta-Analyses extensions for Scoping Reviews (PRISMA-ScR).^(11)^ The research protocol was registered on the Open Science Framework (https://osf.io/tj8mw/).

### Eligibility Criteria

We included randomized controlled trials and observational studies assessing the impact of the Enhanced Recovery After Surgery (ERAS) recommendations on inflammatory markers in patients undergoing laparoscopic surgery. Eligible participants were adults aged 18 years or older who underwent any type of laparoscopic surgery and had one or more of the following inflammatory biomarkers measured: C-reactive protein (CRP), white blood cell count (WBC), immunoglobulins (IgG and IgA), tumor necrosis factor (TNF), total protein (TP), cortisol, among others.

The intervention study groups will be considered those adopting any series of measures aimed at optimizing and accelerating recovery in the perioperative period, while the control group will be considered the population with traditional perioperative care. Both nomenclatures will be considered for the intervention group, both ERAS and Fast-track will be accepted.

### Data Sources and Search Strategy

The literature search was conducted using PubMed (MEDLINE), Scopus, Embase, Web of Science, and the Cochrane Controlled Register of Trials (CENTRAL). We also searched for unpublished studies and gray literature through manual searches of reference lists of included articles. The initial search was performed in January 2024, with a follow-up search in February 2024. Searches employed combinations of MeSH terms and their synonyms including “Enhanced Recovery After Surgery,” “Inflammation,” and “Laparoscopy.” Our search strategies, adapted for each database, are detailed in Appendices I and II. No restrictions were placed on language or publication date.

### Study Selection

Three reviewers (VGVG, LFGP, CDAB) independently screened titles and abstracts using a standardized screening protocol. Full texts of potentially relevant studies were retrieved, and their details uploaded into Rayyan® (Qatar Computing Research Institute, Doha, Qatar).^(12)^ Disagreements among reviewers were resolved through discussion or consultation with a third party (JEGP, LFRF).

### Data Extraction and Risk of Bias Assessment

Data were extracted independently by the same four reviewers using a specially developed form. Extracted information included publication year, country, study type, surgical type, population characteristics, ERAS recommendations details, inflammatory markers, and pertinent findings. Authors were contacted to resolve data discrepancies or clarify missing details.

Risk of bias was assessed using the Guyatt-modified Cochrane approach for randomized trials ^(13, 14)^ and the Morgan approach for non-randomized studies. ^(15)^ The criteria for randomized trials included adequacy of random sequence generation, allocation concealment, blinding (of investigators, patients, data collectors, statisticians, outcome assessors), completeness of outcome data, and absence of selective reporting. A threshold of less than 10% total loss to follow-up was considered low risk. Non-randomized studies were assessed for eligibility criteria, outcome and exposure measurement accuracy, confounder control, and follow-up adequacy.

### Data Analysis and Presentation

Data were synthesized and displayed in two tables highlighting the characteristics of the ERAS recommendations and the inflammatory markers assessed. Details such as publication year, country, study type, surgical type, ERAS details, and outcomes were tabulated. If the outcomes found and summarized in the results are amenable to quantitative analysis, they will be analyzed by means of a meta-analysis, divided into different outcomes.

## RESULTS

### Search Results and Study Selection

Our systematic database search initially identified 64 studies. After removing 9 duplicates, 55 records were screened by title and abstract, with 43 subsequently excluded. Full texts of the remaining 12 studies were evaluated for eligibility, resulting in 5 further exclusions. Ultimately, 7 studies met our inclusion criteria. The selection process is depicted in the PRISMA flowchart (Figure 1).

**Figure 1.**
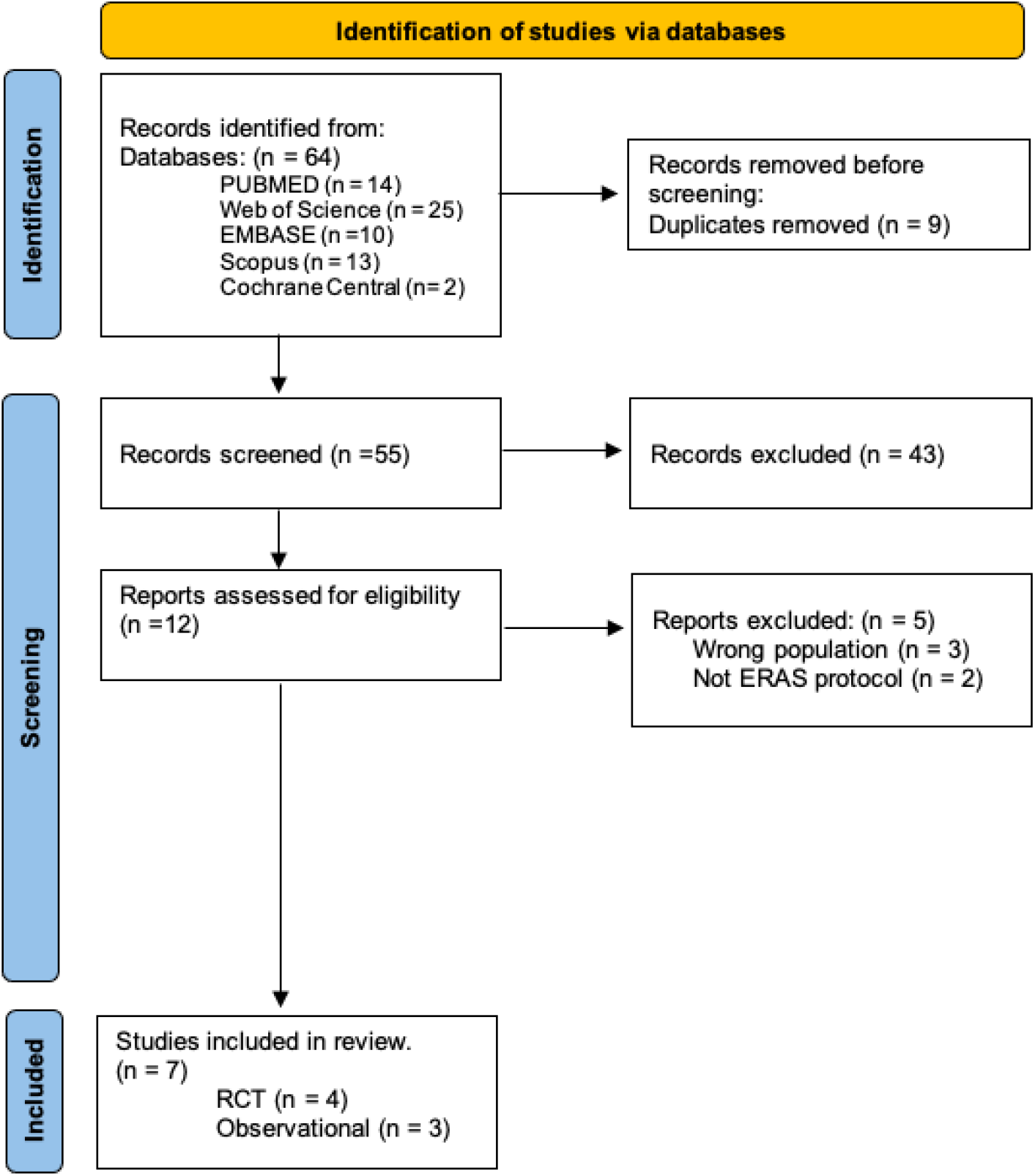
PRISMA Flowchart.

### Characteristics of Included Studies

The included studies were published between 2012 and 2022. Four of these were randomized clinical trials, and three were non-randomized observational studies (Table 1). The studies involved both male and female participants, with the ERAS group having a mean age of 59.7 years-old compared to 49.7 years-old in the control group. Geographically, four studies were conducted in China ^(16-19)^, one in Italy^(20)^, and one in South Korea ^(21)^ (Table 2). The surgical interventions examined included colorectal surgeries^(16-18, 20, 21)^, gastrectomy^(19)^, and gynecological oncological surgery^(22)^.

**Table 1.**
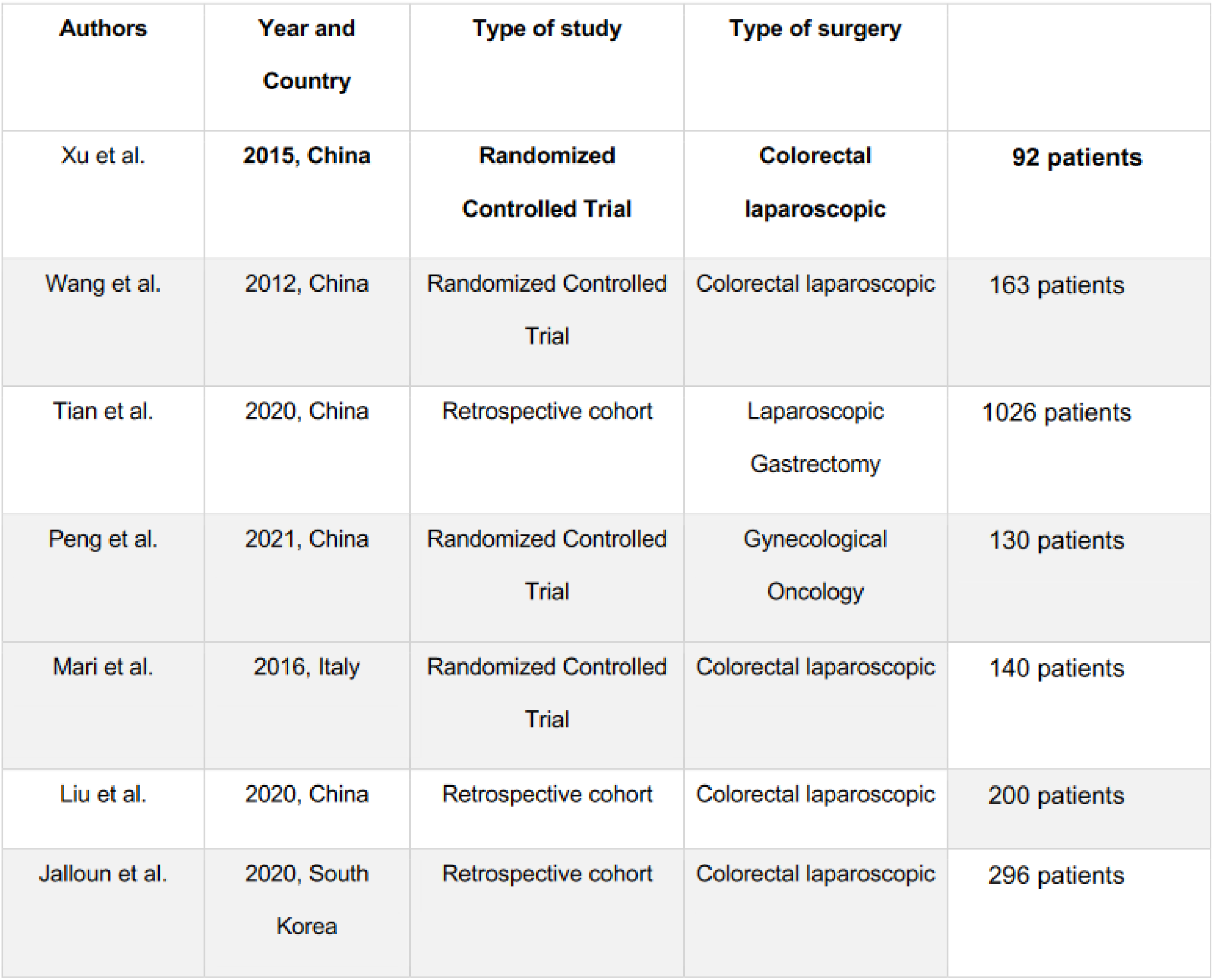
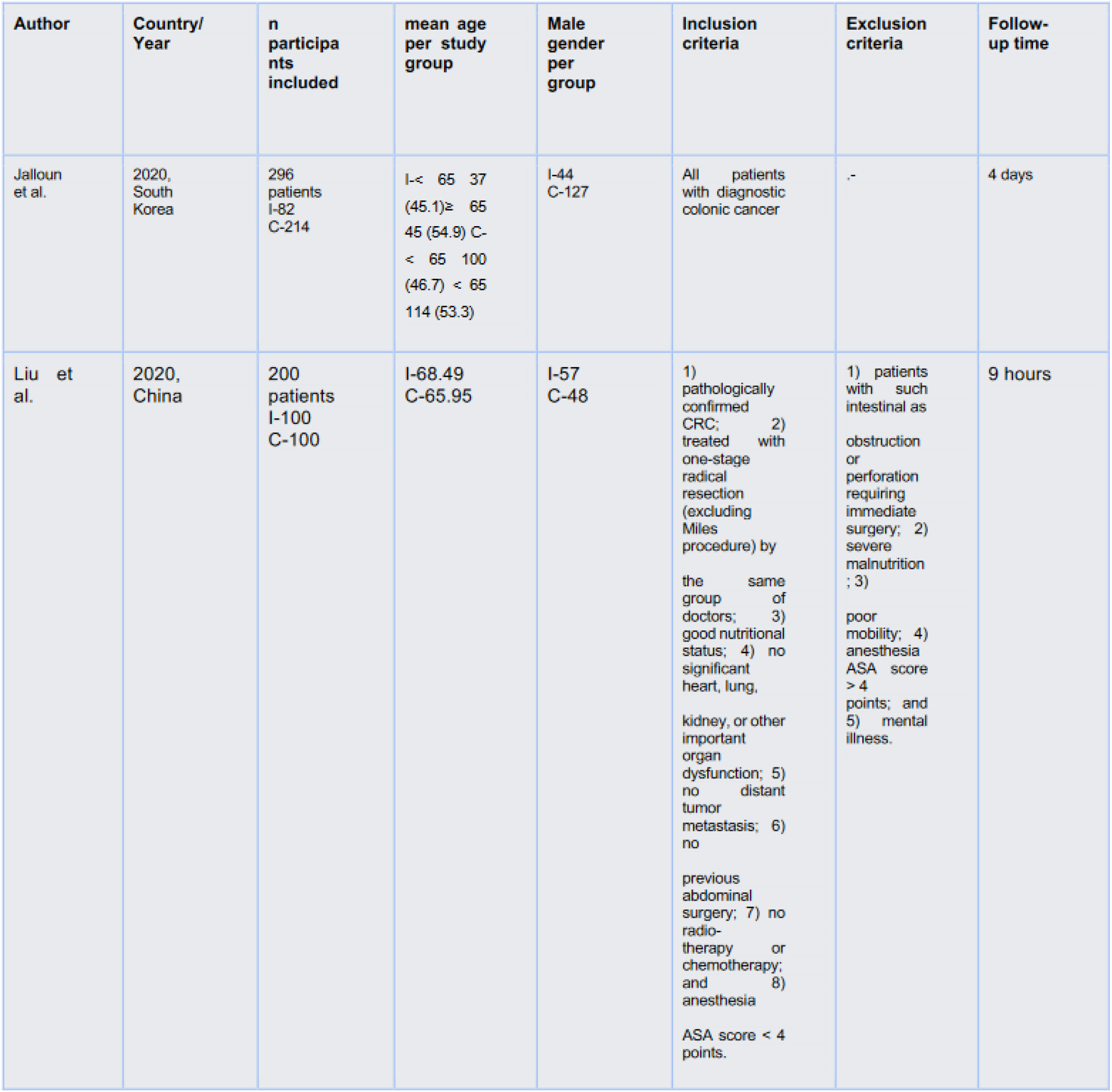
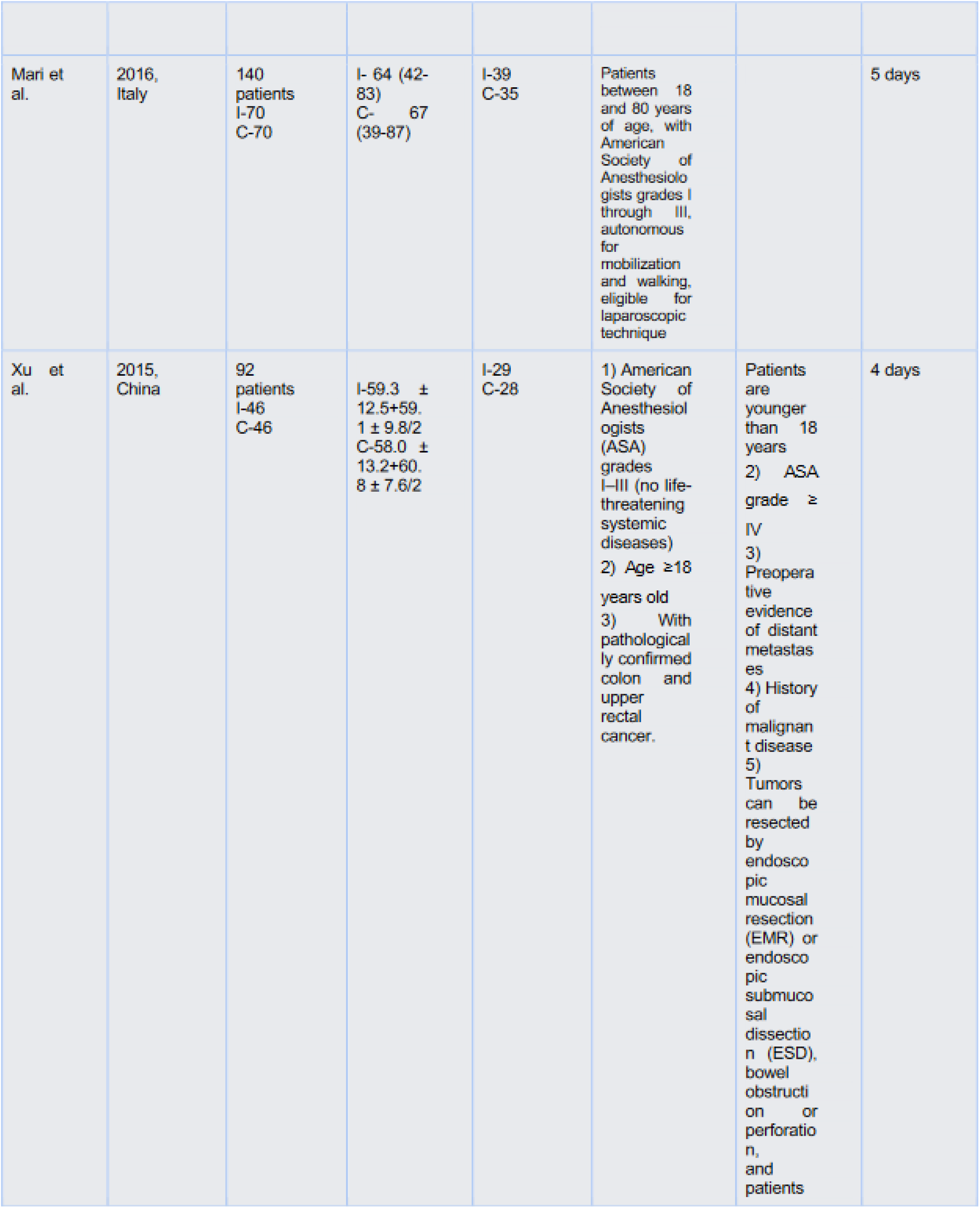
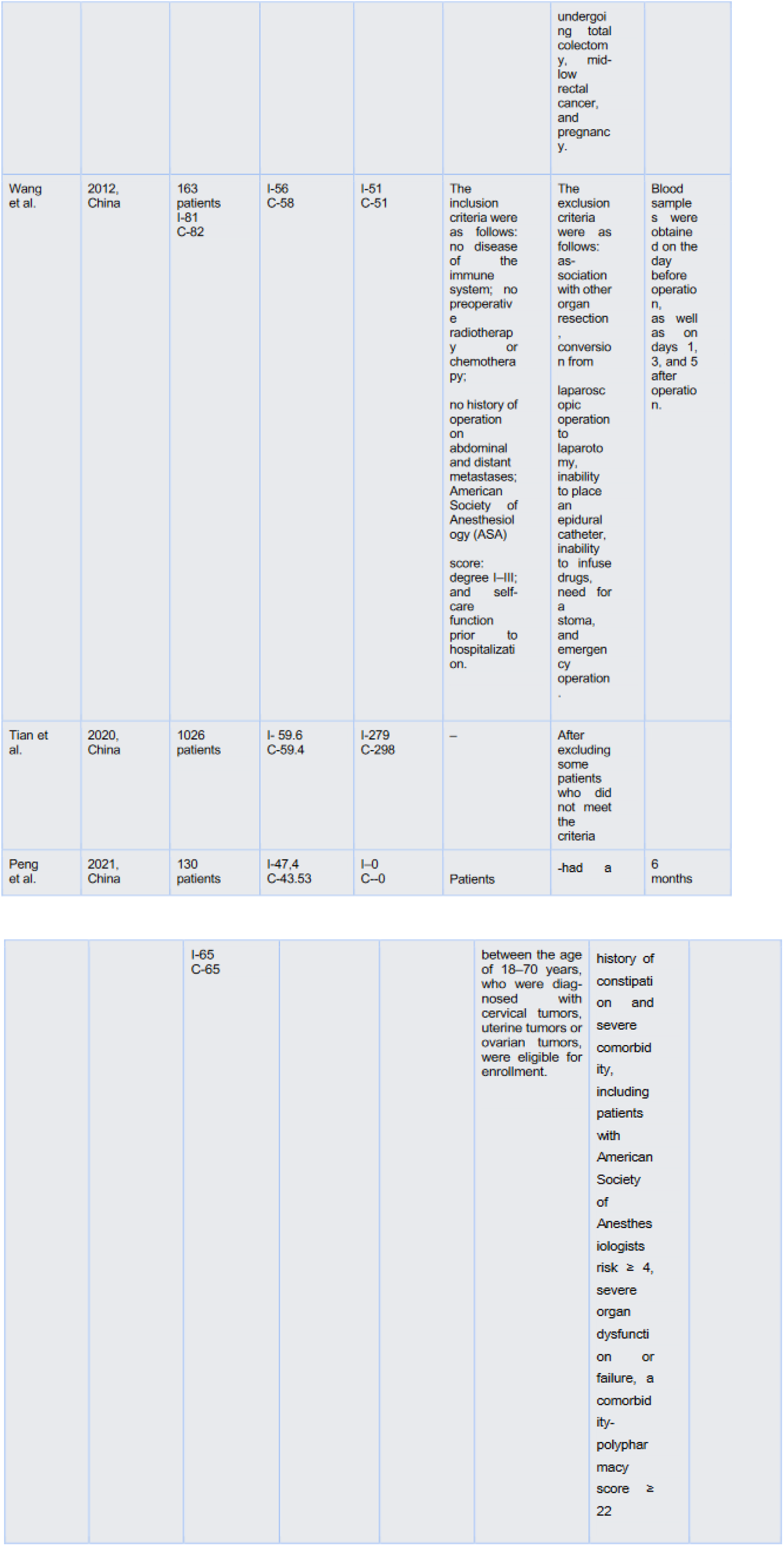
Study characteristics according to population and type of publication.

**Table 2.**
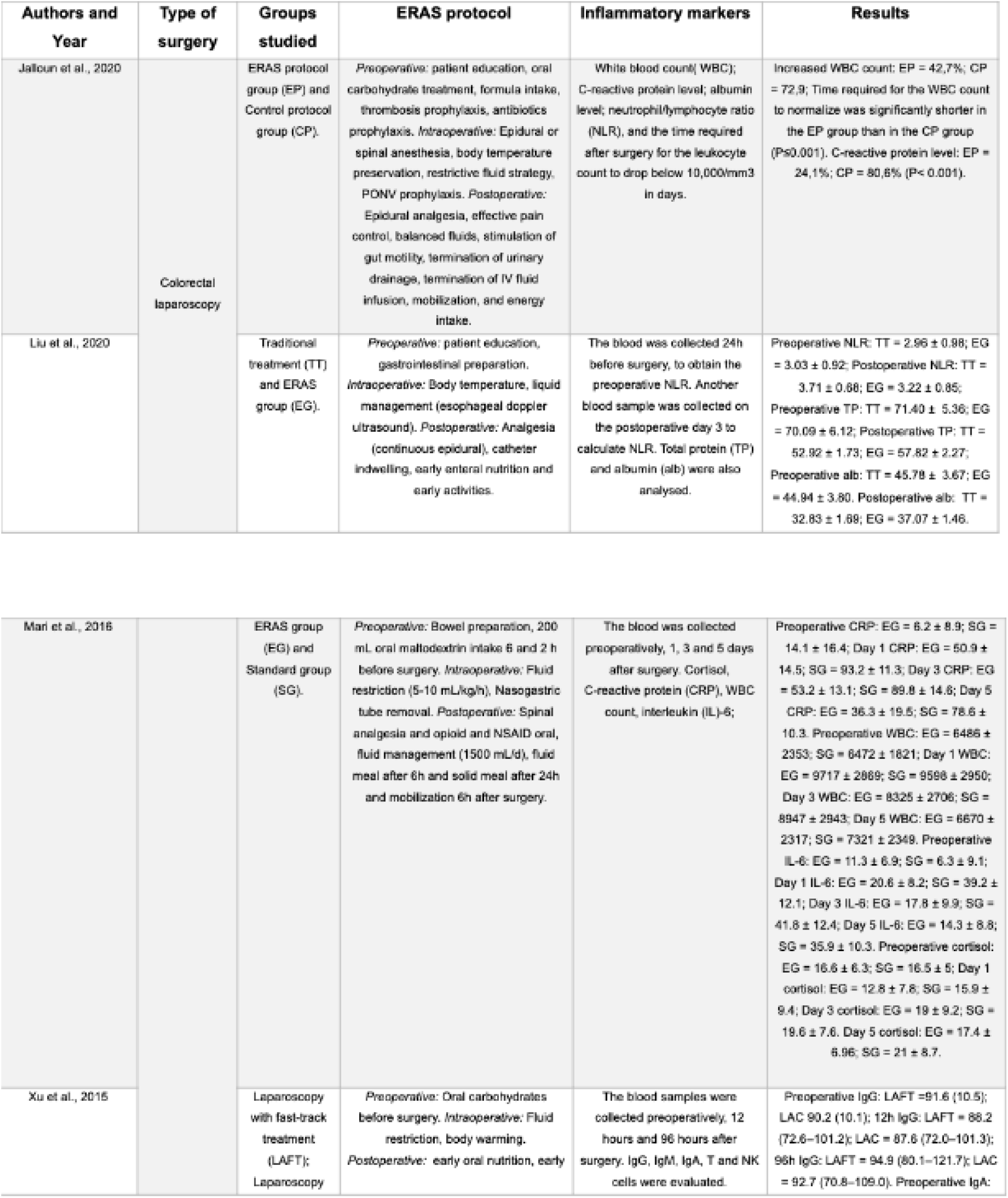
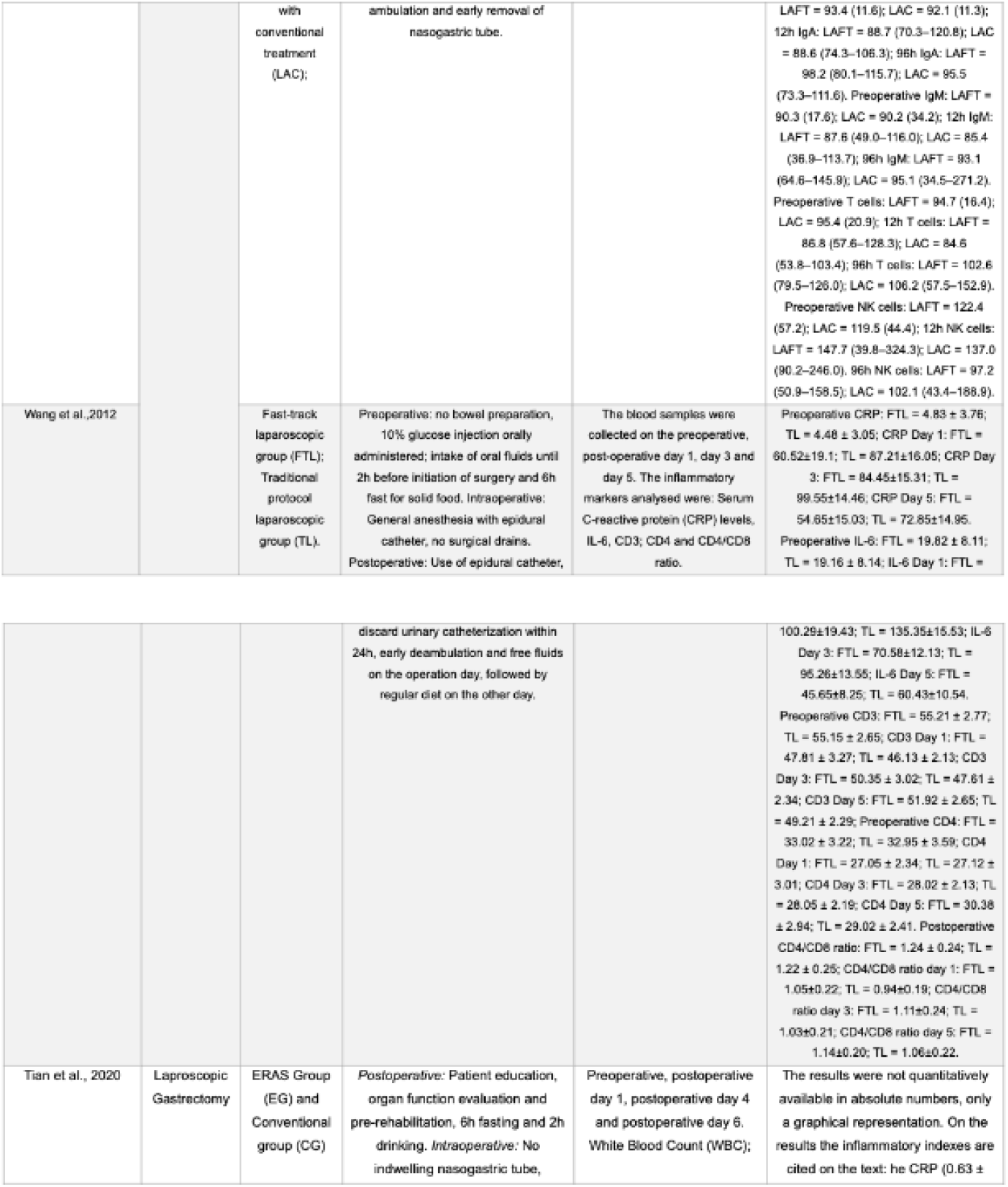
Study characteristics related to population and setting.

### ERAS Recommendations Characteristics

The ERAS recommendations varied but commonly included preoperative patient education, a six-hour fasting period, and carbohydrate-rich liquids up to two hours before surgery. Intraoperative measures focused on fluid restriction, body temperature maintenance, and multimodal anesthetic strategies, including epidural anesthesia in four studies. Postoperative care emphasized early mobilization across all studies.

### Sampling and Inflammatory Markers

Blood samples were collected at various times: six studies ^(16-20, 22)^ during the intraoperative period, one study ^(17)^ 12 hours post-surgery, 4 studies ^(18-21)^ collected on the first postoperative day, 3 studies ^(16, 18, 20)^ collected on the 3rd postoperative day, 2 studies ^(17, 19)^ collected on the 4th postoperative day and 2 studies^(18, 20)^ on the 5th postoperative day, and Tian et al. ^(19)^ was the only study to analyze samples on the 6th postoperative day. Inflammatory markers analyzed included white blood count (WBC) ^(19-22)^, C-reactive protein (CRP)^(18-21)^, albumin^(16, 21)^, neutrophil/lymphocyte ratio (NLR) ^(16, 21, 22)^, total protein (TP) ^(16)^, cortisol ^(20)^, interleukin-6 (IL-6) ^(18, 20)^, immunoglobulin (Ig) G, IgM, IgA^(17)^, T lymphocytes ^(17, 18)^, natural killer (NK) cells ^(17)^, procalcitonin^(19)^, and platelet count^(22)^ (Table 3).

**Table 3.** Assessment of the ERAS protocol and the inflammatory markers analyzed.

| Author/<br>Years | Was the randomization sequence adequately generated? | Was allocation adequately concealed? | Was there blinding of participants? | Was there blinding of the caregiver? | Was there blinding of data collectors? | Was there blinding of statisticians? | Was there blinding of outcome assessors? | Was loss to follow-up (missing outcome data) infrequent?* | Are reports of the study free of suggestion of selective outcome reporting? | Was the study apparently free of other problems that could put it at a risk of bias? |
| --- | --- | --- | --- | --- | --- | --- | --- | --- | --- | --- |
| Xu et al. 2015 | Definitely yes | Probably yes | Probably not | Definitely not | Probably yes | Probably yes | Definitely yes | Definitely not | Probably yes | Definitely yes |
| Wang et al. 2012 | Definitely yes | Probably yes | Probably not | Definitely not | Probably yes | Probably yes | Definitely yes | Definitely not | Probably yes | Definitely yes |
| Peng et al. 2021 | Definitely yes | Probably yes | Probably not | Definitely not | Probably yes | Probably yes | Definitely yes | Definitely not | Probably yes | Definitely yes |
| Mari et al. 2016 | Definitely yes | Probably yes | Probably not | Definitely not | Probably yes | Probably yes | Definitely yes | Definitely not | Probably yes | Definitely yes |

### Risk of Bias in Studies

Randomized studies showed adequate random sequence generation and allocation concealment. Blinding of participants was achieved, but not for surgical staff due to the nature of the procedures. We did not consider this at high risk of bias because those outcomes cannot be influenced by the participants. Blinding of caregivers was considered at high risk due to the impossibility to hide surgical technique from them. Blinding of data collectors, statisticians, and outcome assessors were considered at low risk of bias in all studies. There were no studies reporting total loss to follow-up above the 10% threshold nor above 5% between groups. Therefore, loss to follow-up was considered as low risk of bias (Table 4).

**Table 4.** Risk of bias randomized studies.

| Author<br>year | Do exposed individuals represent the general population? | Was certainty of exposure adequate? | Was selection of non-exposed cohort adequate? | Demonstration that outcome of interest was not present at start of study? | Comparability of cohorts (age) | Comparability of cohorts (other controlled factors) | Assessment of Outcome | Adequate follow-up time | Loss to follow-up |
| --- | --- | --- | --- | --- | --- | --- | --- | --- | --- |
| Categories | SELECTION (1 POINT PER QUESTION) |  |  |  | COMPARABILITY (1 POINT PER QUESTION) |  | OUTCOMES (1 POINT PER QUESTION) |  |  |
| Tian, 2020 | yes | yes | no | yes | yes | yes | yes | no | yes |
| Liu, 2020 | yes | yes | yes | yes | yes | yes | yes | no | no |
| Jalloun, 2020 | yes | yes | yes | yes | yes | probably yes | yes | yes | yes |

Non-randomized studies showed critical. Bias due to confounding was considered critical in two studies ^(19, 21)^, and considered to be serious in one study^(19)^, because they did not correct the adequate non-exposed cohort groups for confounding factors. Bias in selection of participants was considered to be serious in all observational studies ^(16, 19, 21)^, because selection was offered, and not encompassing all the patients. Bias in classification of exposures was considered moderate in three studies^(16, 19, 21)^ because information was self-reported. Bias due to missing data was considered serious in all studies^(16, 19, 21)^ due to the design of the studies (Table 5 and 6).

**Table 5.**
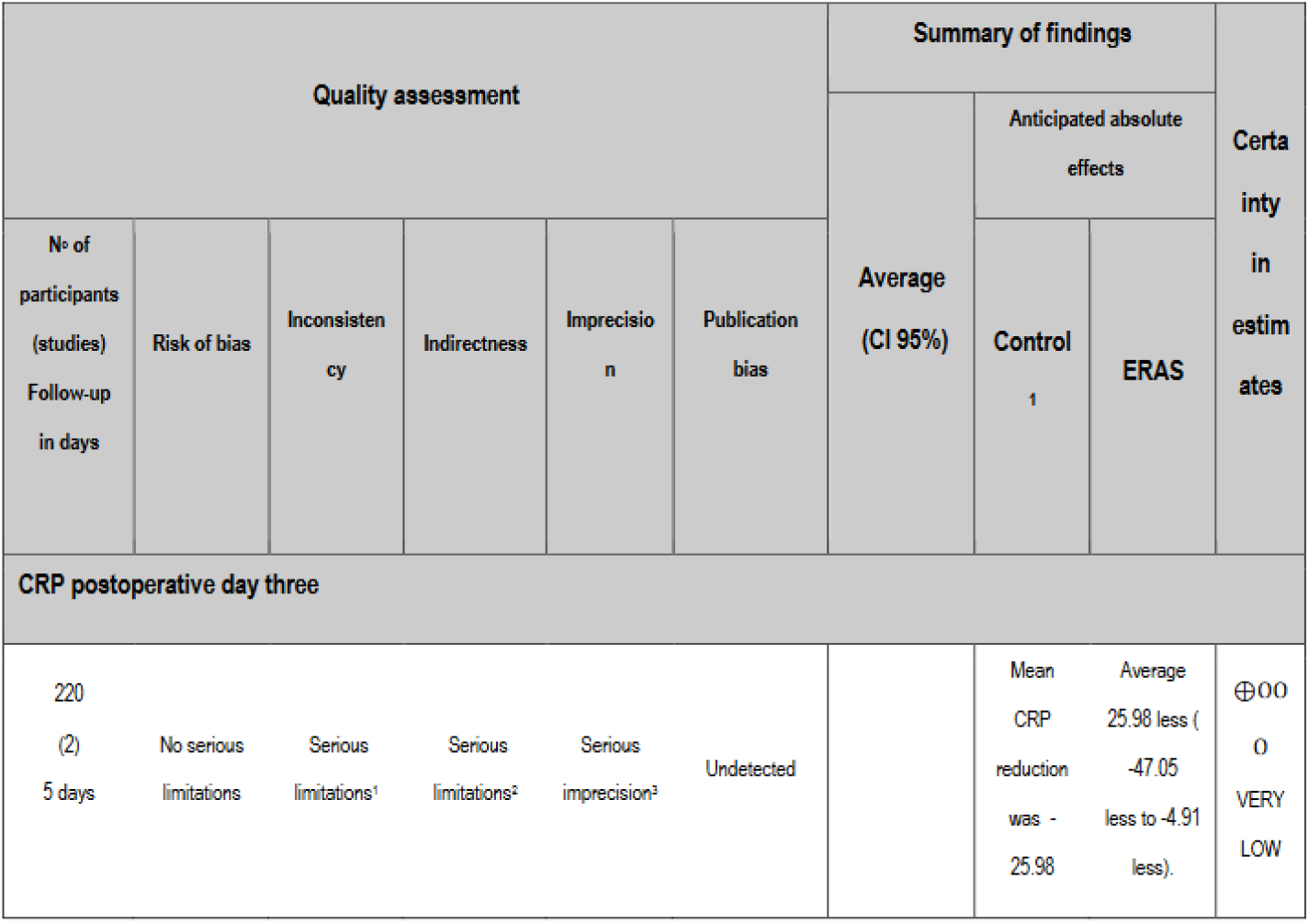
Risk of bias non-randomized studies.

**Table 6.**
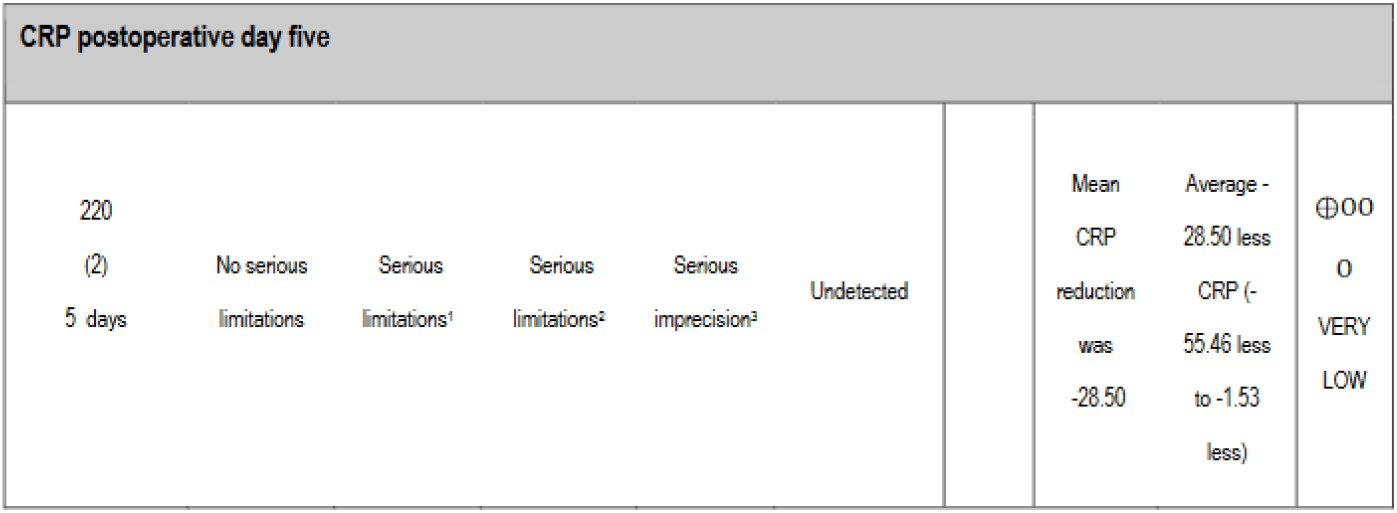
Assessment of GRADE.

1. There was a serious limitation related to inconsistency (12 > 50%).
2. There was a serious limitation related to indirectness (not a clinical outcome)
3. There was a serious limitation related to imprecision (rated down twice due to low number of events and wide confidence intervals).

### Outcomes

*C-reactive protein (CRP):* has been investigated in two observational studies ^(19, 21)^ and two RCTs ^(18, 20)^. The studies included 1,322 patients in the retrospective cohorts^(19, 21)^ and 303 patients in two RCTs ^(18, 20)^. It found significantly lower levels of CRP in ERAS group when compared with standard on postoperative day (POD) 1, 3, and 5. All retrospective studies found lower plasma concentrations of CRP after laparoscopy when compared to open surgery. Jelloun et al. ^(21)^, 2020, including 296 patients, found an increase in CPR count of 24.1% in the ERAS group and an increase of 80.6% in the control group (p < 0.001). Tian et al. ^(19)^, 2020, including 1026 patients, found no significant reduction of CRP in the ERAS group compared to the control group. Mari et al. ^(20)^, 2016, including 140 patients, found that ERAS protocol significantly reduced the rise slope of CRP on postoperative days, 1, 3, and 5 (p<0,05) compared to the control group. Results from two RCTs, including 220 participants, found a significant reduction on the rising slope of C-reactive protein on postoperative day three in the ERAS group compared to the control group (MD -25.98, 95%CI -47.05, -4.91; p=0.02; I^2^=96%). Certainty of evidence was considered very low due to imprecision (wide confidence interval and low number of patients), inconsistency (from high heterogeneity) and indirectness (Figure 2).

**Figure 2.**
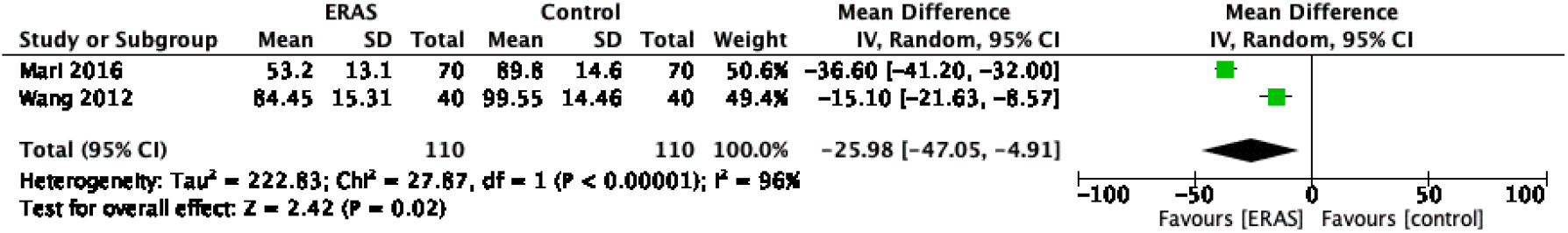
Comparison of CRP on postoperative days three between ERAS and control groups.

Results from two RCTs^(18, 20)^, including 220 participants, found a significant reduction on the rising slope of C-reactive protein on postoperative day five in the ERAS group compared to the control group (MD -28.50 95% CI -55.46, -1.53; p=0.04; I^2^=98%). Certainty of evidence was considered very low due to imprecision (wide confidence interval and low number of patients), inconsistency (from high heterogeneity) and indirectness (Figure 3).

**Figure 3.**
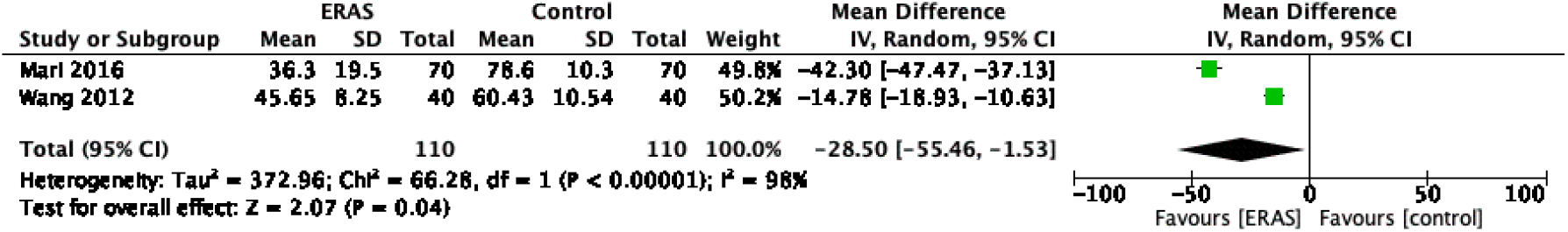
Comparison of CRP on postoperative days five between ERAS and control groups.

*White blood cell count (WBC):* two observational^(19, 21)^ and two RCT ^(20, 22)^ analyzed this inflammatory marker. They found significant reduction of WBC in the ERAS group compared to control on POD 1,3, and 5. Jaloun et al.^(21)^, 2020, including 296 patients, found an increase on WBC count of 42.7% in the ERAS group and an increase of 72.9% in the control group. Time required for the WBC count to normalize was significantly shorter in the ERAS group than in the control group (p ≤ 0.001). Tian et al. ^(19)^, 2020, including 1,026 patients. Although results were not quantitatively available in absolute numbers, a graphic representation suggests that WBC did not increase as much in the ERAS group as it did in the control. Both randomized control trials by Peng et al. ^(22)^, including 130 patients, and Mari et al. ^(20)^, including 140 patients, found no significant reduction of WBC in the ERAS group compared to the control group.

*Interleukin-6 (IL-6):* Two RCTs ^(18, 20)^ evaluated the impact of the ERAS protocol on patients undergoing laparoscopic surgery and the IL-6 marker. A total population of 303 patients, both studies found significantly lower levels of IL-6 on POD 3 after ERAS protocol groups versus standard groups.

## DISCUSSION

This scoping review evaluates the impact of Enhanced Recovery After Surgery (ERAS) recommendation measures during the perioperative period on inflammatory markers in patients undergoing laparoscopic surgery. It aims to elucidate consistent findings, recognize knowledge gaps, and suggest directions for future research on both existing and novel inflammatory markers.

The initial studies following the conceptualization of ERAS by Kehlet et al.^(6)^ focused predominantly on morbidity, mortality, and surgical complications, rather than on the quantitative assessment of inflammatory markers. It took 13 years from the inception of this concept for the first study analyzing the influence of ERAS on inflammatory markers in laparoscopic surgeries to be published.

Laparoscopic colorectal surgery was the most examined procedure, covered in five of the seven included studies. These studies collectively analyzed a total of 2,047 patients, with 891 undergoing colorectal procedures. The largest study, by Tian et al.^(19)^, involved 1,026 patients and examined laparoscopic gastrectomy.

White blood cell count, and C-reactive protein were the most frequently assessed markers, each studied in four investigations. Notably, critical inflammatory markers such as tumor necrosis factor-alpha and alpha 1-acid glycoprotein were absent from the studies reviewed, representing a significant gap in the literature. Markers such as neutrophil gelatinase-associated lipocalin (NGAL) and N-terminal pro B-type natriuretic peptide (NT-proBNP), which indicate organ damage, were also not evaluated.

All reviewed studies reported benefits of the ERAS recommendations over standard care, notably in reducing the surgical stress response, as evidenced by lower levels of IL-6, CRP, and WBCs. Moreover, a trend towards fewer postoperative complications was observed in ERAS patients, although statistical significance was achieved in only one study.

Our findings support the literature showing a beneficial impact of ERAS recommendations, particularly noted in significant reductions of CRP levels on postoperative days three and five (Figures 2 and 3). However, the overall certainty of this evidence remains very low due to issues with precision, consistency, and directness.

This review’s strengths lie in its methodical approach, including a comprehensive search, systematic selection, and rigorous bias assessment, independently replicated by multiple reviewers. Additionally, the GRADE approach was employed to enhance the reliability of evidence evaluation. Conversely, the primary limitations stem from high variability in study outcomes, insufficient blinding of surgical teams, and the reliance on a limited number of small-scale studies, which collectively restrict the precision and applicability of the findings.

This scoping review highlights the need for additional randomized controlled trials (RCTs) to more precisely determine the effects and validate the impact of Enhanced Recovery After Surgery (ERAS) recommendations on inflammatory markers in laparoscopic surgery. The preliminary findings suggest that the implementation of ERAS guidelines may significantly reduce inflammatory markers, potentially leading to enhanced recovery outcomes for patients undergoing laparoscopic procedures. However, due to the limited number of studies and their small sample sizes, the certainty of these findings remains very low, emphasizing the importance of further high-quality research to provide more definitive evidence and refine clinical practice guidelines.

## Data Availability

All data produced in the present work are contained in the manuscript

